# Pathogen genomics and One Health: a scoping review of current practices in zoonotic disease research

**DOI:** 10.1101/2024.02.05.24302264

**Authors:** Stefano Catalano, Francesca Battelli, Zoumana I Traore, Jayna Raghwani, Christina L Faust, Claire J Standley

## Abstract

Whole-genome sequencing has revolutionised the field of infectious disease surveillance, enabling near real-time detection of pathogens and tracking how infections may spread. We performed a scoping review of studies that have applied genomic epidemiology to zoonotic disease transmission across One Health domains (human, animal, and environment). We identified 114 records published between 2005 and 2022 which reported original multi-domain genomic data of zoonotic pathogens integrated into phylogenetic models. Most studies investigated bacterial pathogens, highlighting key knowledge gaps for other zoonotic agents, particularly arboviruses. Sampling and sequencing efforts vastly varied across domains: the median number and range of pathogen genomes analysed were highest for humans (23; 1-29,586) and lowest for the environment domain (13; 1-956). Infectious disease genomics was leveraged to track zoonotic disease outbreaks and cross-domain transmission, to enhance pathogen surveillance, and to disentangle evolutionary dynamics driving lineage diversification and virulence. Our study informs effective study design for future genomic applications to multi-domain and cross-species transmission of zoonoses, with the potential to identify key infection sources and inform interventions for local and global health security.

## INTRODUCTION

Zoonoses are infectious diseases that can transmit naturally from animals to humans (https://www.who.int/news-room/fact-sheets/detail/zoonoses). Approximately 60% of emerging infections are caused by zoonotic pathogens (Jones *et al*, 2008; Becker *et al*, 2019). While it is difficult to ascertain the true impact of these infections, zoonoses often pose a double burden by affecting human health while also being of veterinary concern, often leading to significant economic losses in animal production (Noguera *et al*, 2022). Managing and preventing zoonotic disease outbreaks requires interdisciplinary approaches and expertise from a diversity of fields (WHO, 2019; UNEP & ILRI, 2020; Kock & Caceres-Escobar, 2022). A One Health approach encourages close collaboration among different disciplines and sectors to recognise that human and animal health are interdependent and intricately connected to the health of the environment, or ecosystem, in which they coexist. The checklist for One Health epidemiological reporting of evidence (COHERE) was developed to guide the integration of knowledge across these three domains (i.e., human, animal, and environment) when designing and implementing interventions (Davis *et al*, 2017). However, investigating multiple domains within a One Health context remains complex and necessitates overcoming many hurdles, including diagnostics capacity and supply chain limitations, policy and funding support, and meaningful equal participation from a wide variety of stakeholders (Munyua *et al*, 2019; Ribeiro *et al*, 2019).

At the beginning of the 21^st^ century, the advent of the first high-throughput sequencing platforms ushered in the next-generation sequencing era, driving rapid and financially accessible sequencing of entire genomes (Goodwin *et al*, 2016). The application of genomics to the transmission dynamics of infectious diseases enabled estimates of fine-scale epidemiological processes within a relatively short time span (Biek *et al*, 2015; Didelot *et al*, 2017). The West African Ebola virus disease epidemic and the coronavirus disease 2019 (COVID-19) pandemic were powerful reminders of the impact of zoonoses on human populations and have underscored the importance of real-time surveillance to elucidate transmission pathways (Gire *et al*, 2014; Keita *et al*, 2021; Urban *et al*, 2023). In this context, efforts to predict onward transmission events after initial pathogen spillover in susceptible host populations and across One Health domains have increased dramatically after the emergence of SARS-CoV-2 (Carrasco-Hernandez *et al*, 2017; Wasik *et al*, 2019; UNEP & ILRI, 2020; Hopkins *et al*, 2022; Kock & Caceres-Escobar, 2022). Applying genomic tools to One Health research offers the exciting prospect of reconstructing transmission events via genomic epidemiology and phylodynamics. When combined with qualitative and quantitative epidemiological data, these relatively recent approaches can help uncover the origin of an outbreak, transmission routes, and/or potential super-spreading events (Hill *et al*, 2021; Ingle *et al*, 2021). Furthermore, zoonotic disease mitigation cannot bypass the identification of competent disease reservoirs, which benefits from progress in sampling, diagnostics, sequencing, and modelling techniques (Viana *et al*, 2014; Becker *et al*, 2020). In the context of public health, diagnostic centres and health agencies are rapidly adopting these techniques, particularly to investigate foodborne outbreaks and to track the epidemic potential of influenza viruses (Armstrong *et al*, 2019).

Leveraging genomic data and modelling approaches within fieldwork settings and across different domains remains logistically and analytically complex. As a result, the degree to which these approaches have been used to investigate zoonotic disease transmission is poorly understood. In this scoping review, we scanned published literature and extracted data to the finest attainable methodological, spatiotemporal, and phylogenetic level of detail to characterise sampling strategies, genomic approaches, and evolutionary models applied to zoonoses within One Health initiatives. Our objectives were to identify studies investigating cross-domain zoonotic pathogen transmission at the human-animal and/or human-environment interfaces using genomic epidemiology. Our study aimed to characterise and categorise genomic applications to One Health research and, in doing so, to highlight current practices and knowledge gaps for informing future studies.

## RESULTS

### Record screening

The literature search yielded 2,094 and 1,637 results for PubMed^®^ and Web of Science™, respectively, for a total of 3,731 results. Automatic de-duplication produced a list of 3,030 records and was followed by manual screening, which removed a further 36 records. The list of records subjected to title/abstract primary screening contained 2,992 results. The primary screening led to 272 conflicting decisions out of 2,992 records (90.9% agreement rate). After conflict resolution, a total of 2,723 records were excluded while 269 articles were included for full-text screening. Among the excluded records, a substantial amount focused on SARS-CoV-2 (n=164). The secondary screening based on full texts led to the exclusion of 155 records (57.6%) while 114 (42.4%) were included for data extraction (Appendix Fig S2).

Overall, most studies targeted bacterial pathogens (83.3% (n=95)) whereas viral and parasitic organisms were less represented (Table 3). The 114 studies included for data extraction covered 36 different families, genera, or species of infectious disease agents (i.e., 23 bacteria, seven RNA viruses, three protozoa, one fungus, one nematode, and one DNA virus). The distribution of the studies amongst pathogen taxa was highly skewed with 17 infectious disease agents represented by only one study. Of the 95 studies on bacterial pathogens, almost half (48.4% (n=46)) focused on one of three species: *Salmonella enterica, Escherichia coli,* or *Staphyloccocus aureus*. RNA viruses were represented by 13 studies (11.4%), which focused on seven genera (i.e., *Alphainfluenzavirus*, *Flavivirus*, *Kobuvirus*, *Orthohantavirus*, *Orthonairovirus*, *Phlebovirus*, and *Rotavirus*) but four of these were only represented by one study.

### Geographic, temporal, and motivation trends of genomics applied to One Health

Our dataset included 92 records originating from 33 different countries, whereas 22 publications collected and/or analysed multiple specimens from at least three different countries. Most records (74.6% (n=85)) analysed specimens exclusively from upper-middle/high income countries whereas only 10.5% (n=12) focused solely on low/lower-middle income countries (sampling and sequencing of specimens from both income status groups was included by 13.2% (n=15) of the studies). The People’s Republic of China was the most represented country when studies focused their sampling efforts on a single country (n=15), followed by the USA (n=9) and Australia (n=9).

We observed an increasing trend in the average number of published studies annually between 2011 and 2019, followed by a decline in last three years (2020-2022 but our search stopped in September 2022) (Fig 1). For all included studies, the average time lag between sampling end date and publication year was 3.8 years (median 3; range 0-21). Most records (79.8% (n=91)) had a time lag of five years or less, whereas fewer records (9.6% (n=11) and 4.4% (n=5)) had six to 10 years and 11 to 21 years as time lags, respectively (Fig 2B).

**Figure 1.**
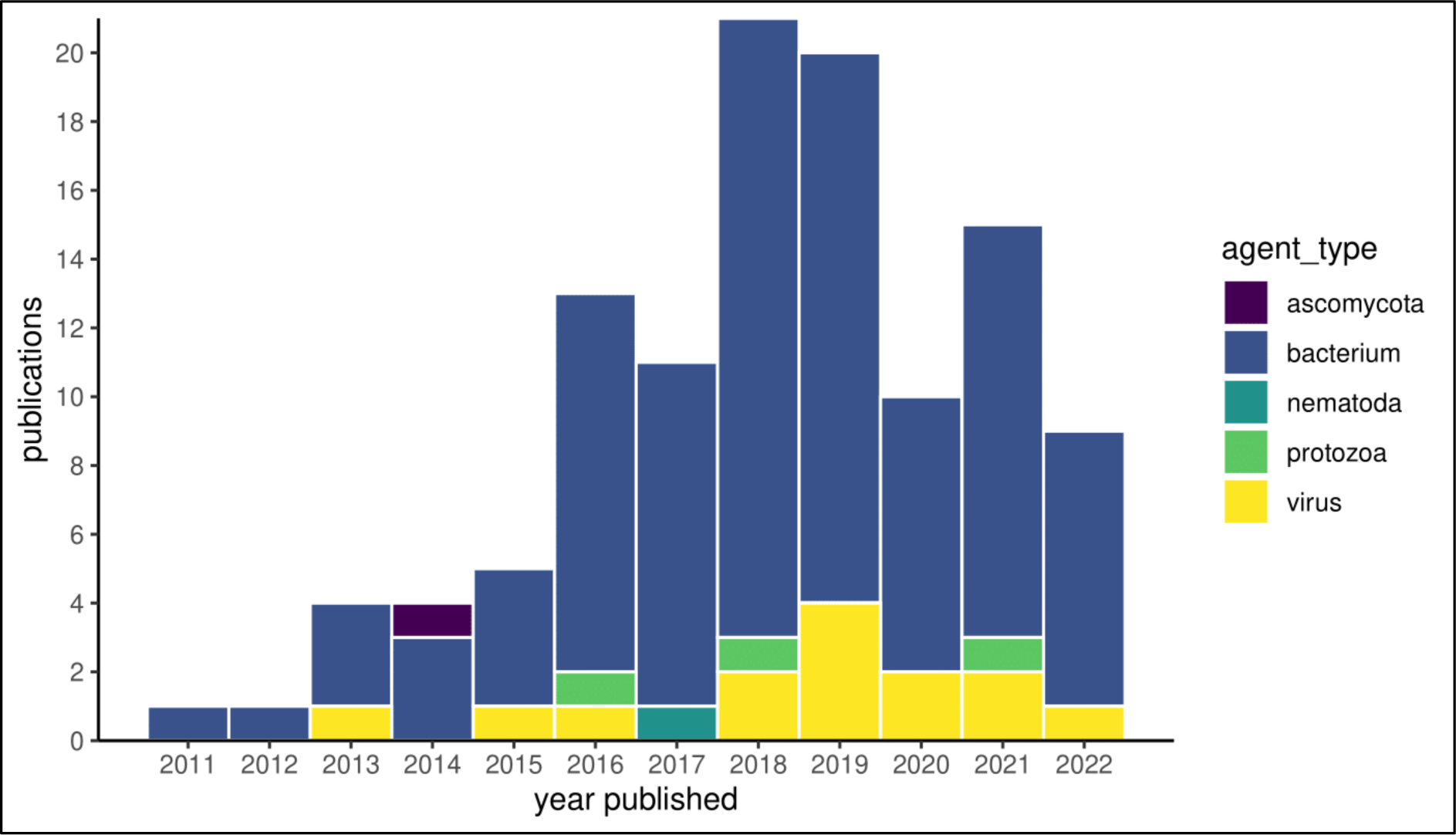
Publication year and zoonotic agent type. Cumulative studies including genomic data and addressing at least two One Health domains (i.e., human, animal, and environment) shaded by the focal zoonotic pathogen(s) for each of the 114 records included in this study.

**Figure 2.**
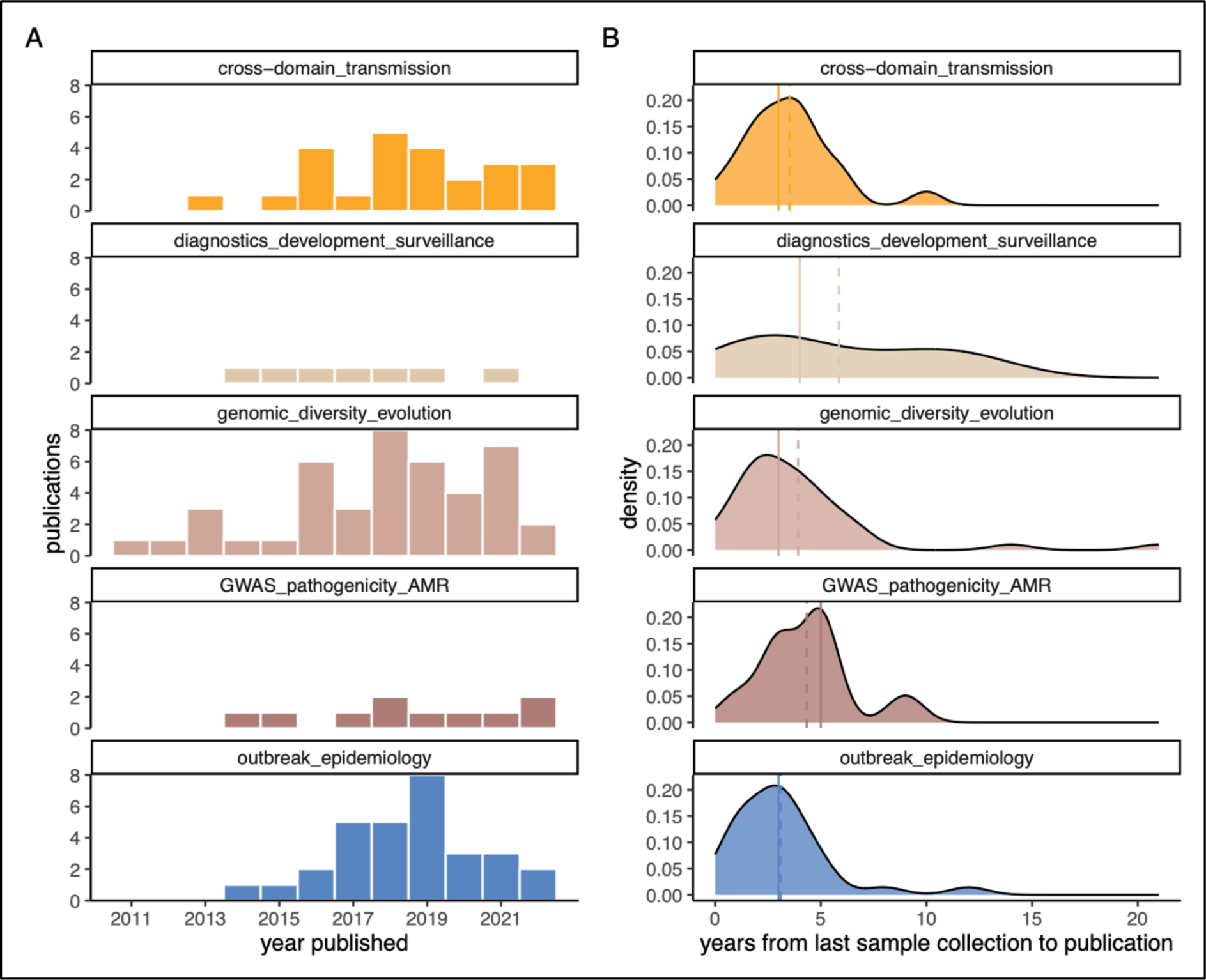
Overarching aim of each study. A Histograms representing the number of publications for each overarching study aim (i.e., cross-domain transmission, diagnostics development and surveillance, genomic diversity and evolution, genome-wide association studies (GWAS) including antimicrobial resistance (AMR), and outbreak epidemiology investigations) and year of publication. B Density plots displaying the delay between the most recently collected specimen and year of publication (the dotted and solid lines are the mean and median time lags, respectively).

The overarching aim of each study was classified into five different categories. We assigned the following motivations: cross-domain transmission, diagnostics development and surveillance, genomic diversity and evolution, genome-wide association studies including antimicrobial resistance (GWAS-AMR), and outbreak epidemiology investigations. The most common aim was genomic diversity and evolution (37.7% (n=43)), followed by outbreak epidemiology investigations (26.3% (n=30)). Diagnostics development and surveillance was the least explored aim, with only 6.1% (n=7) of the studies targeting it (Fig 2A and Appendix Table S4). The breakdown into the different study aims showed that outbreak epidemiology investigations had the lowest mean time lag of 3.1 years (median 3; range 0-12), followed by cross-domain transmission with 3.5 years (median 3; range 0-10), genomic diversity and evolution with 3.9 years (median 3; range 0-21), GWAS-AMR with 4.3 years (median 5; range 1-9), and diagnostics development and surveillance with 5.9 years (median 4; range 1-12) (Fig 2B).

The application of different sequencing systems over time shows a consistent delay of at least two years between the technology’s commercialisation and the first scientific publication(s) applying it to One Health studies (Fig 3). The only exception is PacBio^®^ RS II, an instrument which appeared in scientific publications soon after its commercial release. Illumina^®^ sequencing platforms represented the most widely used systems, particularly MiSeq and HiSeq (respectively employed as the primary platform in 33.3% (n=38) and 29.8% (n=34) of the studies). Long-read sequencing by PacBio^®^ RS II and/or Oxford Nanopore Technologies MinION was used by 14.0% (n=16) of the records as either the main instrument or as support to short-read sequencing of bacteria (n=14), Zika virus (n=1), and *Plasmodium* spp. (n=1).

**Figure 3.**
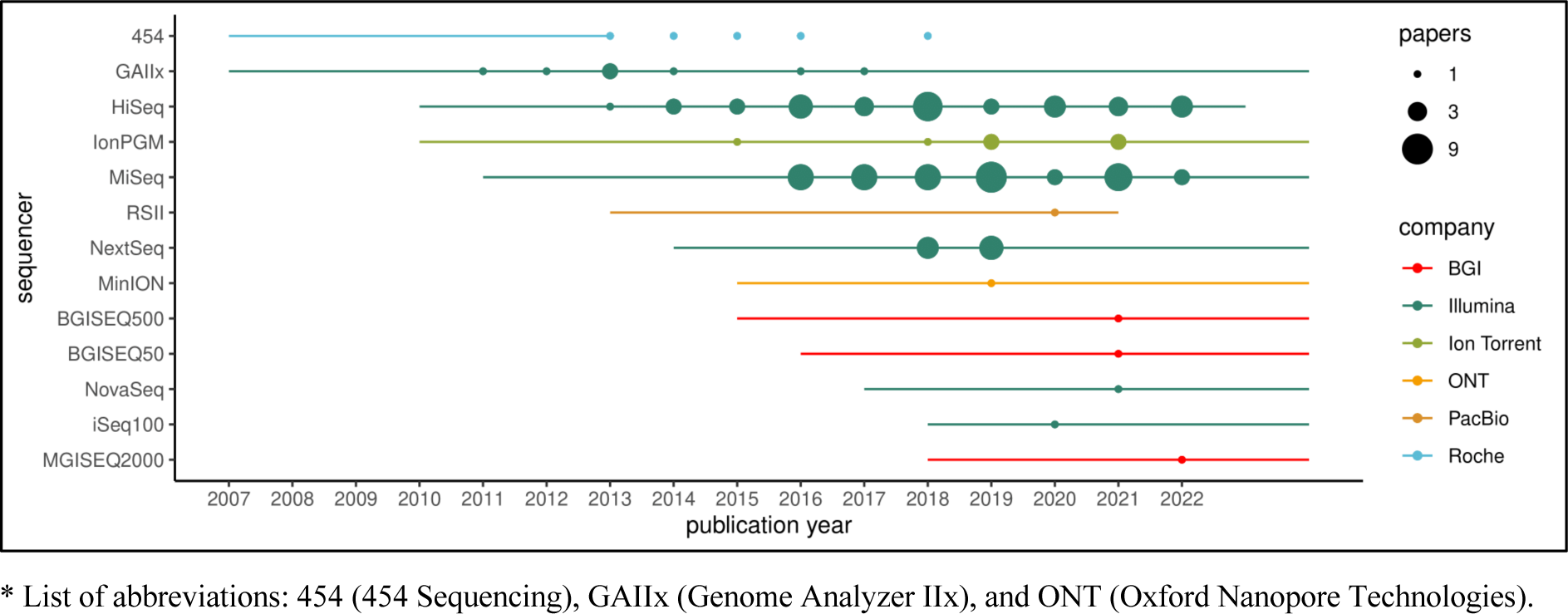
Sequencing platforms and frequency of use in One Health studies over time. Information on the lifespan of each instrument was collated until 2023. For each technology, the line shows the year of commercialisation and discontinuation when available. The point size reflects the year of publication for each of the 114 records included in this study and the specific technologies used for genomic sequencing*. * List of abbreviations: 454 (454 Sequencing), GAIIx (Genome Analyzer IIx), and ONT (Oxford Nanopore Technologies).

### Multi-domain analysis of infectious disease agents

In our dataset, the human, animal, and environment domains were simultaneously investigated by 37.7% (n=43) of the records. Most of these three-domain studies investigated bacterial pathogens (n=39) (of these studies, 23 focused on either *E. coli*, *S. enterica*, or *S. aureus*) whereas few of these three-domain studies focused on RNA viruses (n=3) or protozoa (n=1). Almost two-thirds of the studies analysed and sequenced specimens either at the human-animal (38.6% (n=44)) or human-environment interface (23.7% (n=27)) (Table 1). Studies that focused on zoonotic diseases at the animal-environment interface were excluded during abstract/title and full-text screening.

**Table 1.** List of zoonotic pathogens. Zoonotic pathogens investigated in the 114 studies included for data extraction subdivided by the surveyed One Health domains (human (HUM), animal (ANI), and environment (ENV)). The total number of records for each taxon is reported in parentheses.

| Pathogen |  | One Health domains |  |  |
| --- | --- | --- | --- | --- |
| Classification | Taxon | HUM-ANI-ENV | HUM-ANI | HUM-ENV |
| Gram-negative bacteria | *Salmonella enterica* (22) | 13 | 8 | 1 |
|  | *Escherichia coli* (13) | 8 | 3 | 2 |
|  | *Burkholderia pseudomallei* (5) | 1 | 0 | 4 |
|  | *Campylobacter* spp. (3) | 0 | 3 | 0 |
|  | *Francisella tularensis holartica* (3) | 1 | 1 | 1 |
|  | *Yersinia pestis* (3) | 2 | 0 | 1 |
|  | *Coxiella burnetii* (2) | 1 | 1 | 0 |
|  | *Vibrio parahaemolyticus* (2) | 0 | 0 | 2 |
|  | *Acinetobacter baumannii* (1) | 0 | 0 | 1 |
|  | *Chlamydia psittaci* (1) | 0 | 1 | 0 |
|  | Enterobacteriaceae family (1) | 0 | 0 | 1 |
|  | *Klebsiella pneumoniae* (1) | 0 | 0 | 1 |
|  | *Leptospira* spp. (1) | 1 | 0 | 0 |
|  | *Orientia tsutsugamushi* (1) | 1 | 0 | 0 |
|  | *Rickettsia japonica* (1) | 0 | 0 | 1 |
|  | **Total (60)** | **28** | **17** | **15** |
| Gram-positive bacteria | *Staphylococcus aureus* (11) | 2 | 7 | 2 |
|  | *Listeria monocytogenes* (5) | 2 | 1 | 2 |
|  | *Mycobacterium* spp. (5) | 2 | 3 | 0 |
|  | *Bacillus anthracis* (4) | 3 | 1 | 0 |
|  | *Enterococcus* spp. (3) | 1 | 1 | 1 |
|  | *Streptococcus* spp. (3) | 0 | 3 | 0 |
|  | *Bacillus cereus* (2) | 1 | 0 | 1 |
|  | *Clostridium* spp. (2) | 0 | 1 | 1 |
|  | **Total (35)** | **11** | **17** | **7** |
| Single-stranded RNA viruses | *Flavivirus* genus (3) | 1 | 0 | 2 |
|  | *Influenza A virus* (3) | 1 | 1 | 1 |
|  | *Orthohantavirus* genus (3) | 0 | 3 | 0 |
|  | CCHF *orthonairovirus* (1) | 1 | 0 | 0 |
|  | *Kobuvirus* genus (1) | 0 | 1 | 0 |
|  | *Phlebovirus* genus (1) | 0 | 0 | 1 |
|  | **Total (12)** | **3** | **5** | **4** |
| Others | *Babesia microti* (1) | 0 | 0 | 1 |
|  | *Cowpox virus* (1) | 0 | 1 | 0 |
|  | *Giardia duodenalis* (1) | 1 | 0 | 0 |
|  | *Plasmodium* spp. (1) | 0 | 1 | 0 |
|  | *Rotavirus A* (1) | 0 | 1 | 0 |
|  | *Sporothrix* spp. (1) | 0 | 1 | 0 |
|  | *Strongyloides stercoralis* (1) | 0 | 1 | 0 |
|  | **Total (7)** | **1** | **5** | **1** |

Sample sizes were highly variable across domains. The median number of specimens analysed and sequenced was highest in humans (23; range 1-29,586) and closely followed by animals (21; range 1-2,004). Environmental samples had the smallest median sample size and range (13; range 1-956). Of all publications that included environmental specimens (n=70), 52.9% (n=37) analysed and sequenced abiotic samples. In contrast, a focus on vectors was only included by 22.9% (n=16) of these studies (Fig 4A). The median number of specimens was highest from arachnids (14; range 1-175), followed by abiotic sources (12; range 1-559), biotic sources (5.5; range 1-956), and insects (2.5; range 1-18). Among studies that sequenced specimens from the animal domain (n=87), sub-category representation was highest for specimens from livestock (63.2% (n=55)) followed by wildlife (36.8% (n=32)), poultry (34.5% (n=30)), and pets (24.1% (n=21)) (Fig 4A). Of these 87 studies, 66.7% (n=58) included specimens from multiple non-human vertebrate species. The median number of specimens was highest from both livestock (13; range 1-760) and poultry (13; range 1-1,244), followed by wildlife (7; range 1-116) and pets (3; range 1-18).

**Figure 4.**
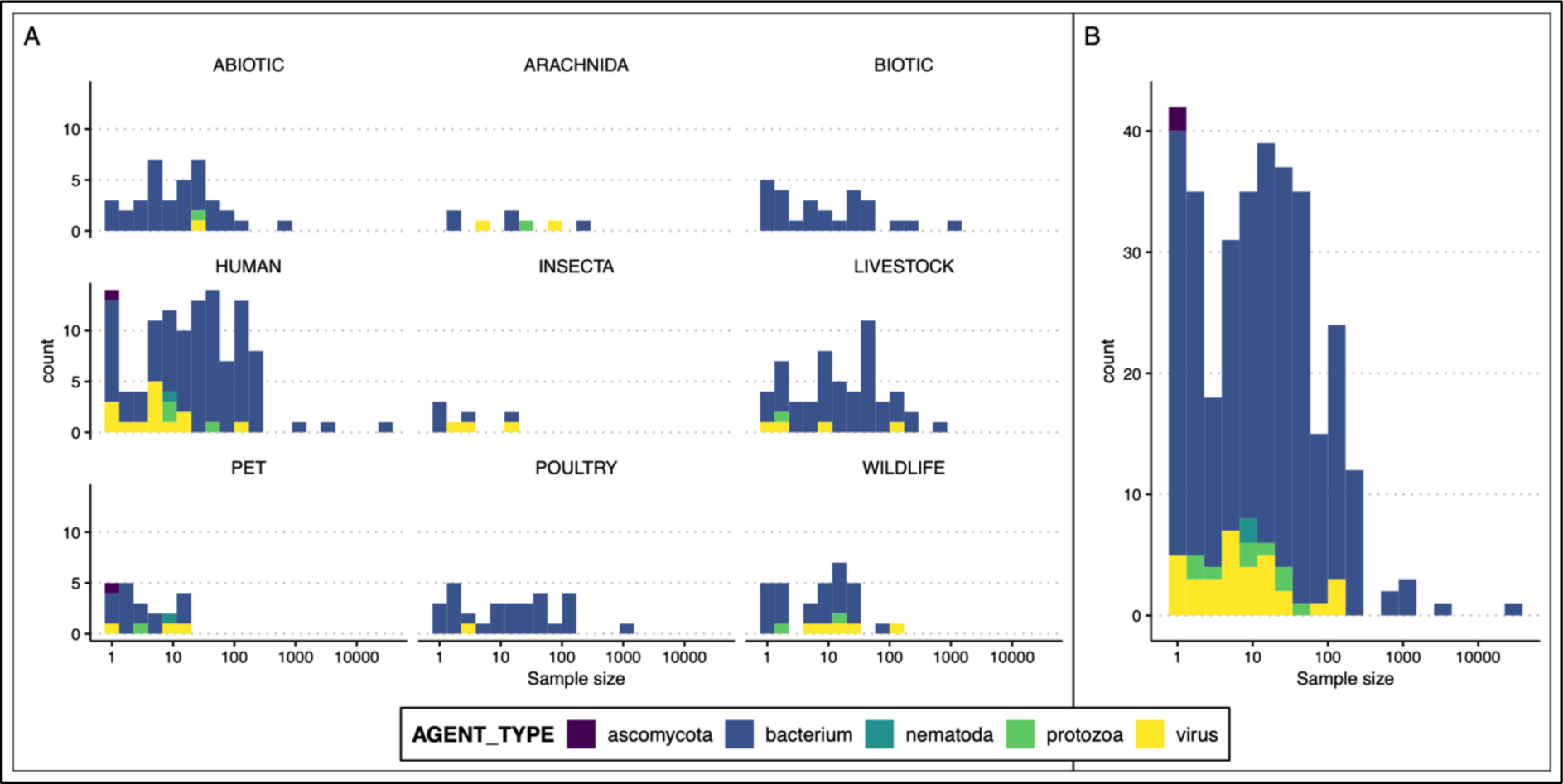
Counts of specimens collected (or analysed) and sequenced by each study shaded by the type of infectious disease agents. A Histograms representing the sample size reported by each publication (x-axis) and the number of studies (y-axis) for each One Health domain (i.e., human, animal (subdivided into livestock, pet, poultry, and wildlife), and environment (subdivided into abiotic, Arachnida, biotic, and Insecta)) from which the analysed specimens originated. B Histogram representing the sample size reported by each publication (x-axis) and the number of studies (y-axis).

We employed a generalised linear model to investigate how total sample size of each record was influenced by different variables (i.e., pathogen type, biocontainment level, sequencing platform, sampling geographic origin, income status stratification, overarching aim of the study, number of surveyed domains, and year of publication). Models were all over-dispersed and, therefore, we could not construct a model that sufficiently described sample size distributions even when using negative binomial families. As a result, the total sample sizes were not predicted by study aim (Appendix Fig S3), sequencing platform, biocontainment level as defined by Health and Safety Executive (https://www.hse.gov.uk/pubns/misc208.htm), number of surveyed domains, sampling geographic origin, income status stratification, and year of publication. Only publications focusing on bacterial pathogens had significantly larger sample sizes across all domains (Fig 4B) (n=95 studies; median 68.5; range 2-31,292) when compared to viruses (n=14 studies; median 8; range 6-219), or parasites (i.e., fungi, helminths, and protozoa) (n=5 studies; median 18; range 2-89).

### Analytical trends of genomics applied to One Health

We extracted data on genome assembly software/pipelines from each paper and identified 23 unique toolkits used by the 114 records (Appendix Table S5). SPAdes (Prjibelski *et al*, 2020) was the most widely used assembly toolkit, implemented by 28.9% (n=33) of the records. This is a freely available software supporting a wide array of data types and pipelines, leading it to being the most selected tool for both *de novo* assembly and mapping to a reference genome. CLC Genomics Workbench (QIAGEN, Aarhus), a licensed software suite for integrated genomic data analytics, was also frequently used, almost exclusively for *de novo* assembly of short-read data (14.0% (n=16) of the studies). In contrast, Burrows-Wheeler Aligner (Li & Durbin, 2009) was often utilised for mapping reads to a reference genome (10.5% (n=12) of the studies). Most studies utilized non-commercial, freely available software/pipelines to assemble or align genomic data (71.9% (n=82)). However, applied coding pipelines were rarely made available alongside publications (Appendix Fig S4).

Furthermore, we extracted details of statistical approaches and tools employed for building phylogenetic trees using sequencing data from multi-domain pathogens. Overall, maximum likelihood estimation of phylogenetic relatedness was the most utilised inference method (72.8% (n=83 studies)) amongst those applied (i.e., neighbor-joining, minimum spanning network, maximum parsimony, maximum likelihood, and Bayesian inference). Bayesian inference was only applied by 19.3% (n=22) of the records, although it was the dominant method to estimate species divergence timescales (used by 18 out of the 19 studies which enforced molecular clocks) (Fig 5). Phylogenetic model validation using a combination of approaches was undertaken by a total of 30 studies, of which more than one-third (n=12) applied both maximum likelihood and Bayesian inference, one-third (n=10) applied both neighbor-joining and maximum likelihood, and one-sixth (n=5) validated phylogenetic models using three different inference methods.

**Figure 5.**
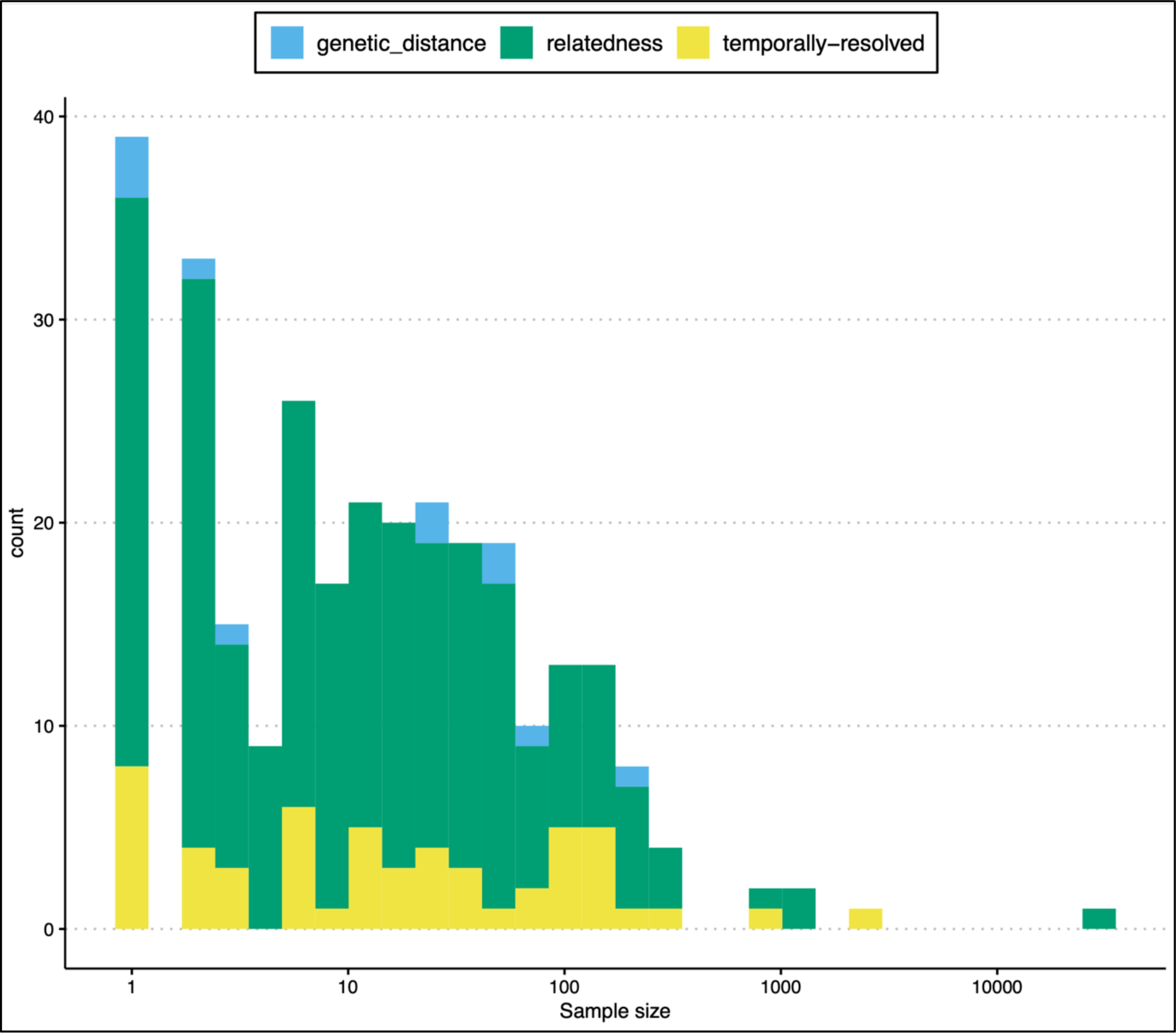
Phylogenetic models of zoonotic pathogen genomes in One Health. Number of records (y-axis) and reported sample sizes (x-axis) shaded by the main output of the evolutionary model implemented by each study. Genetic distance (blue) defined the application of neighbor-joining or minimum spanning networks only. Relatedness (green) represented studies inferring evolutionary relationships using maximum likelihood, maximum parsimony, or Bayesian inference. Temporally resolved (yellow) indicated more computationally intensive models enforcing molecular clocks.

A variety of software was used to build phylogenetic trees (Appendix Table S6). RAxML (Stamatakis, 2014), a freely available program for maximum likelihood estimation, was most utilised (28.1% (n=32) of the studies), followed by BEAST and BEAST 2 (Suchard *et al*, 2018; Bouckaert *et al*, 2019) for Bayesian evolutionary inference (16.7% (n=19)) and MEGA software (15.8% (n=18)). Most studies used non-commercial, freely available software to build phylogenetic trees (85.1% (n=97)) and deposited raw reads or curated genomic data into public archives (86.8% (n=99)). Publications originating from either low/lower-middle income countries or upper-middle/high income countries did not show specific assortment with open science practices (Appendix Fig S4).

## DISCUSSION

Diseases rarely occur in closed, isolated populations and failing to account for such complexity may result in models unable to replicate underlying transmission pathways. Nevertheless, the depth of information provided by genomic data already plays a critical role in pathogen characterisation, disease surveillance, and preventative strategies, while genome databases continue to grow vastly and are becoming extensive big data resources in infectious disease research (e.g., Carroll *et al*, 2018; Li *et al*, 2021; Stockdale *et al*, 2022). Our scoping review provides epidemiological and analytical insight into genomic studies investigating transmission and diversity of zoonotic pathogens across multiple One Health domains. We retrieved 114 studies whose sampling and research effort was heterogenous across infectious agents, domains, geographies, sequencing technologies, bioinformatic toolkits, and phylogenetic modelling. Below, we discuss such heterogeneities and highlight opportunities for addressing current knowledge gaps.

Our work observed that whole-genome sequencing of bacterial pathogens is a major focus of genomic studies of zoonotic diseases across domains (95 out of 114 publications). In contrast, viruses and pathogens with complex transmission pathways and multi-host life cycles (i.e., helminths and protozoa) were poorly represented. Genomic advances have transformed both fundamental research and public health surveillance of bacterial pathogens, particularly foodborne agents (e.g., *E. coli* and *S. enterica*) which have become pilot organisms for routine whole-genome sequencing and genomic epidemiology studies (Gilchrist *et al*, 2015; Allard *et al*, 2016). Genomic studies paired with phylogenetic approaches can reconstruct recent epidemic histories, providing insight into potential sources and timing of pathogen emergence, transmission events and pathways that may be contextualised temporally and spatially (Faria *et al*, 2017; Ridenour *et al*, 2021; Forde *et al*, 2022). To date, genomic epidemiology, and phylodynamics in particular, seemed to be more restricted to single-stranded RNA viruses owing to their small, rapidly mutating genomes, which require lower sequencing effort to detect contemporary phylogenetic signal (Fountain-Jones *et al*, 2018; Ingle *et al*, 2021). The underrepresentation of viruses in One Health zoonotic studies may reflect the difficulty in isolating and amplifying their genomes in multi-domain contexts. However, historical trends demonstrate that spillover events of zoonotic viruses are increasingly frequent and leading to more severe epidemics (Meadows *et al*, 2023). Therefore, evolutionary epidemiology of viral communities is crucial to establish spatial, temporal, and environmental traits which may support cross-species transmission forecast and public health risk mitigation (Mutz *et al*, 2022).

More than one-third of the records which we included for data extraction explored zoonotic transmission across all One Health domains by integrating human, animal, and environmental components. This is an exciting finding since zoonotic disease control is increasingly recognised as a complex, multi-factorial issue requiring concerted responses across different sectors, including public health, environmental management, veterinary medicine, and agriculture (Davis *et al*, 2017; Standley *et al*, 2023). Nevertheless, investigating all One Health domains may not always be a priority, or even necessary, for certain pathogens or research questions. For example, infectious disease organisms such as *Campylobacter* bacteria and *Orthohantavirus* single-stranded RNA viruses are characterised by relatively short environmental persistence (Bronowski *et al*, 2014; Guterres & de Lemos, 2018), which may lead investigators to disregard the inclusion of environmental specimens. Similarly, the epidemiological role of non-human vertebrate hosts, including their competence in pathogen transmission and spillover mechanisms, remains a work in progress for many infectious disease organisms (Plowright *et al*, 2017; Becker *et al*, 2020). Consequently, One Health research may not feel the urge to investigate potential animal reservoirs of zoonotic diseases until serendipitous findings shed light on their suitability as alternative hosts (McDonald *et al*, 2020; Tomori & Oluwayelu, 2023).

Overall, a lower effort was placed into applying genomic tools to environmental specimens within multi-domain initiatives. Particularly for vector-borne pathogens, we found that a small percentage of publications combined human data with surveys of vectors (i.e., arachnids and insects), while the median number of specimens analysed from insects was the lowest among the sub-categories across all domains. This knowledge gap is striking given that vector-borne zoonotic diseases such as dengue, Rift Valley fever, and West Nile fever are mosquito-borne public health priorities in many regions worldwide. A clearer understanding of the transmission dynamics between animal reservoir, arthropod vector, and human hosts is essential for control strategies and interventions (Plowright *et al*, 2017; Veo *et al*, 2019). Excitingly, metagenomic sequencing of individual or pooled arthropod vectors offers a potential single assay to comprehensively identify vector species, vector-borne pathogens, and animal hosts that define their transmission cycle (Batson *et al*, 2021).

Only 27 out of 114 studies were based in low/lower-middle income countries. Yek *et al* (2022) clearly highlighted the challenges that low/lower-middle income countries face in embracing the so-called genome sequencing revolution, with numerous obstacles that are intrinsic to resource-scarce settings such as access to education and retention of skilled personnel, availability of sequencing platforms, reagents, and maintenance service, financial sustainability, analytical bottlenecks, and access to research initiatives and data. A further obstacle in resource-scarce settings may be implementing genomic surveillance beyond the few centralised hubs that currently exist. Across the African continent, a recent assessment reported that approximately 71% of sequencing systems resides in just five countries, most of them at laboratories with no affiliation to national public health institutes (Inzaule *et al*, 2021). Nevertheless, low/lower-middle income countries are precisely where genomic pathogen surveillance applied to One Health initiatives is more appropriately deployed based on the challenges driven by climate emergencies, land-use change, and zoonotic disease risks which these regions face (Allen *et al*, 2017; Baker *et al*, 2022; Noguera *et al*, 2022).

We noted an increase in the number of published studies over time but a drop between 2020 and 2022, plausibly ascribed to the COVID-19 pandemic which has captured most of the scientific attention in recent years. This is also demonstrated by the high number of records focusing on SARS-CoV-2 which we excluded during the primary screening stage. Furthermore, the social dynamics and preventative measures enforced during COVID-19 may have resulted in a decline of other zoonotic disease outbreaks (Onyeaka *et al*, 2021; Ma *et al*, 2022). In fact, outbreak epidemiology investigation was a principal aim of the publications which we included for data extraction. In contrast, the least explored aim included the development of diagnostics and pathogen surveillance efforts, possibly because this is already a necessary step leading to the more targeted genomic applications which we have evaluated as part of our scoping review.

Our objective was to identify, in a transparent and reproducible manner, relevant records modelling genomic data to track zoonotic pathogen transmission across One Health domains. However, we defined One Health merely using a human-centric perspective on zoonotic disease epidemiology without including other socio-ecological aspects shaping community and ecosystem health (Gallagher *et al*, 2021; Stephen *et al*, 2023). We deliberately chose to limit our screening to studies that included genomic sequencing of specimens from human hosts. Therefore, we omitted the substantial body of work integrating genomics to decipher pathogen transmission among multiple non-human vertebrate hosts and at the animal-environment interface (e.g., Bennett *et al*, 2020; Forde *et al*, 2022).

Since the introduction of next-generation sequencing technology in 2005, the array of high-throughput systems has dramatically evolved, including inherent costs, capabilities, and applications (Goodwin *et al*, 2016; Maljkovic Berry *et al*, 2020). Our study confirmed that Illumina^®^ systems are the most used short-read sequencers, particularly MiSeq which was employed by one-third of the studies as the main sequencing system, confirming previous findings (Goodwin *et al*, 2016; Segerman, 2020). MiSeq remains the most common sequencing platform for infectious disease research and public health, likely owed to its fast runtime, compact benchtop design, and strong user support (Goodwin *et al*, 2016; Maljkovic Berry *et al*, 2020). Other platforms, such as Ion Torrent and MGI sequencers, were used to produce short-read data in our survey but less frequently. For long-read sequencing, PacBio^®^ RS II and/or Oxford Nanopore Technologies MinION were used by 14% of studies as either primary platforms or to supplement short-read sequencers. Most of these publications focused on bacterial pathogens likely due to their highly complex genome structures including long, repetitive, and mobile elements which require validation using long-read data (Wick *et al*, 2023). Such studies integrating multiple sequencing approaches with complementary strengths are a positive example about avoiding potential systemic biases in the produced data (Stoler & Nekrutenko, 2021; Ali *et al*, 2023).

Maximum likelihood clearly emerged as the most utilised inference method, particularly to estimate evolutionary relatedness among isolates, possibly owing to its computational efficiency and robustness while remaining conceptually a relatively simple method (Dhar & Minin, 2016). However, less than one-third of the records combined two to three methods for model validation. Validating phylogenetic models by using different approaches is a fundamental step to ensure adequate realism, robustness, and reproducibility of the outputs in the contexts of deciphering transmission dynamics and forecasting disease outbreaks (Heesterbeek *et al*, 2015; Rees *et al*, 2021). The main reason for the lack of extensive model validation may be attributed to the analyst’s habit in model construction. In our study, evolutionary models that included three domains and/or above-average sample sizes did not necessarily translate to accurate model validation. In some cases, phylogenetic inference was relatively simple for studies including intense sampling efforts (e.g., Tsui *et al*, 2018; Sia *et al*, 2020; Hudson *et al*, 2021). Nevertheless, sampling effort and evolutionary model complexity should not be mistaken for realism; many studies may choose to focus on a lower sampling effort and on a single interface (i.e., human-animal or human-environment) and apply relatively simple phylogenetic methods which allowed them to accurately depict transmission dynamics. For example, Black *et al* (2019) only sequenced a small number of clinical specimens and *Aedes* mosquitoes to specifically estimate pathogen introduction and phylogeographic dynamics of Zika virus during an epidemic in Colombia. Maximum likelihood estimation of temporally resolved phylogenies alone strongly suggested that onward transmission of circulating strains played a far stronger role than multiple introduction events in determining the magnitude of the Zika virus disease epidemic across Colombia. Despite some methodological caveats, including the absence of bootstrap resampling and molecular clock model validation using Bayesian inference, the small sample size and the study’s objective seemed to justify the authors’ decision to opt for a lower complexity of the evolutionary model.

## CONCLUSIONS

Our scoping review identified several key areas for future progress in the application of genomic technologies to infectious disease research across multiple One Health domains. First, the need to integrate multi-domain surveillance of arboviruses and their vectors is clear. The environment as a whole was less considered as a source of zoonotic disease transmission. Second, high-throughput sequencing in low- and middle-income countries remains insufficiently leveraged to study zoonotic diseases across domains. Resource-scarce settings still face numerous challenges in the application of ‘omics technologies but these regions are exactly where the deployment of these tools would greatly benefit pathogen surveillance within One Health initiatives. Finally, we observed a gap in the practice of evolutionary model validation by combining different inference methods. Model validation is a crucial step in establishing that phylogenetic trees depict ancestral divergencies that are reasonably accurate, adequately supported, and reproducible.

To adequately address the threat of emerging pathogens, scientific and public health communities must join forces to harness the power of whole-genome sequencing technologies in One Health and to develop a roadmap for ensuring research capacity strengthening and sustainability (Yek *et al*, 2020; Urban *et al*, 2023). Within One Health, and more in general interdisciplinary research, the migration to whole-genome studies has recently begun and a large expansion is still expected. Admittedly, genomic approaches to One Health require multidisciplinary collaboration among many different fields to adequately interpret data and conclusions (Chakraborty & Barbuddhe, 2021). These collaborations require time and effort to establish, especially between sectors whose research has long remained siloed. However, there is a clear need for more studies exploring the human-animal-environment triad and next-generation sequencing has coalesced into powerful tools for infectious disease detection and epidemiology. In the current climate, the potential for sophisticated evolutionary modelling to inform future surveillance efforts and control strategies is generating significant enthusiasm. Therefore, it is essential that future initiatives will start combining these elements to generate robust models using a One Health approach that can predict outbreak dynamics and changes in disease transmission risks to inform surveillance efforts and public health policy development.

## METHODS

### Search strategy

We followed published guidance on conducting (Munn *et al*, 2018; Foo *et al*, 2021) and reporting (Tricco *et al*, 2018) evidence synthesis. First, we searched PROSPERO database to determine whether our research questions had not been already addressed by a registered review (Moher *et al*, 2015). Then, we queried the following search engines based on their large, multidisciplinary spectrum and their classification as principal resources (Gusenbauer & Haddaway, 2020): PubMed^®^ and Web of Science™ (Web of Science Core Collections selected within the platform). Intentionally, we did not use the Medical Subject Headings (MeSH) database in PubMed^®^ to include any non-indexed article and to restrict our query to the exact search string.

To ensure that the search string had the ability to capture all relevant articles, two authors examined the full text of several studies on zoonotic transmission dynamics (i.e., Yadav *et al*, 2019; Durrant *et al*, 2020; Kim *et al*, 2020; Medkour *et al*, 2020; Gee *et al*, 2021; Zhang *et al*, 2022). This exercise enabled the identification of the terminology applied to our search string and the inclusion criteria, which were then discussed and finalised with other authors (Appendix Table S1). The final search string applied by our study was the following: (ecolog* OR evolution* OR epidemiolog*) AND (“transmission” OR “surveillance”) AND (zoono* OR “disease” OR infect*) AND (“molecular” OR genetic* OR genom* OR metagenom*) AND (phylogen* OR phylodynamic* OR phylogeograph*) AND (“reads” OR librar* OR align* OR polymorph* OR “next generation”) NOT (Sanger OR microsatellite*).

The search was completed within one day on September 27^th^, 2022. Records were exported to EndNote X9.3.3 (Clarivate™, Philadelphia, USA) and combined into a single library. This scoping review followed the Preferred Reporting Items for Systematic reviews and Meta-Analyses extension for Scoping Reviews (PRISMA-ScR) guidelines (Appendix Table S2 adapted from Tricco *et al* (2018)).

### Record screening

Screening of articles was performed using a three-stage process. In the first stage, the library was de-duplicated using EndNote, followed by a visual check of the record list sorted by digital object identifier. In the second stage, two reviewers independently screened titles and abstracts in Rayyan (Ouzzani *et al*, 2016); 100 randomly selected records were initially screened to ensure an agreement rate of at least 80% between reviewers before proceeding with title/abstract review of all records. In the third stage, two reviewers independently screened the full text of each retained article; 10 randomly selected records were first screened to ensure an agreement rate of at least 80% between reviewers before proceeding with full-text review of included records. At each stage, the reviewers followed a decision tree, which was defined by the inclusion and exclusion criteria listed below (Appendix Fig S1). The resolution of any conflicting classifications was addressed by a discussion between reviewers; if needed, the full paper was retrieved and re-screened to resolve the disagreement.

Records that complied with the following inclusion criteria were retained: I) the infectious agent(s) is classified as zoonotic or deemed a potentially emerging zoonotic disease by the publication and/or co-authors of this scoping review (Appendix Table S3 adapted from Rees *et al* (2021)); II) the record includes sampling activities (or handling of specimens for nucleic acid extraction, library preparation, and sequencing) of human hosts in addition to the animal and/or environment One Health domains (in other words, the record includes genomic data produced directly by the study from the human domain in addition to the animal and/or environment One Health domains) (Table 2 adapted from Cavalerie *et al* (2021)); III) genomic data are integrated into evolutionary models of transmission dynamics; and IV) articles’ publication date goes from January 1^st^ 2005 to present (this criterion was based on the commercial release of the first high-throughput sequencing platforms (Goodwin *et al*, 2016; Kulski, 2016)).

**Table 2.**
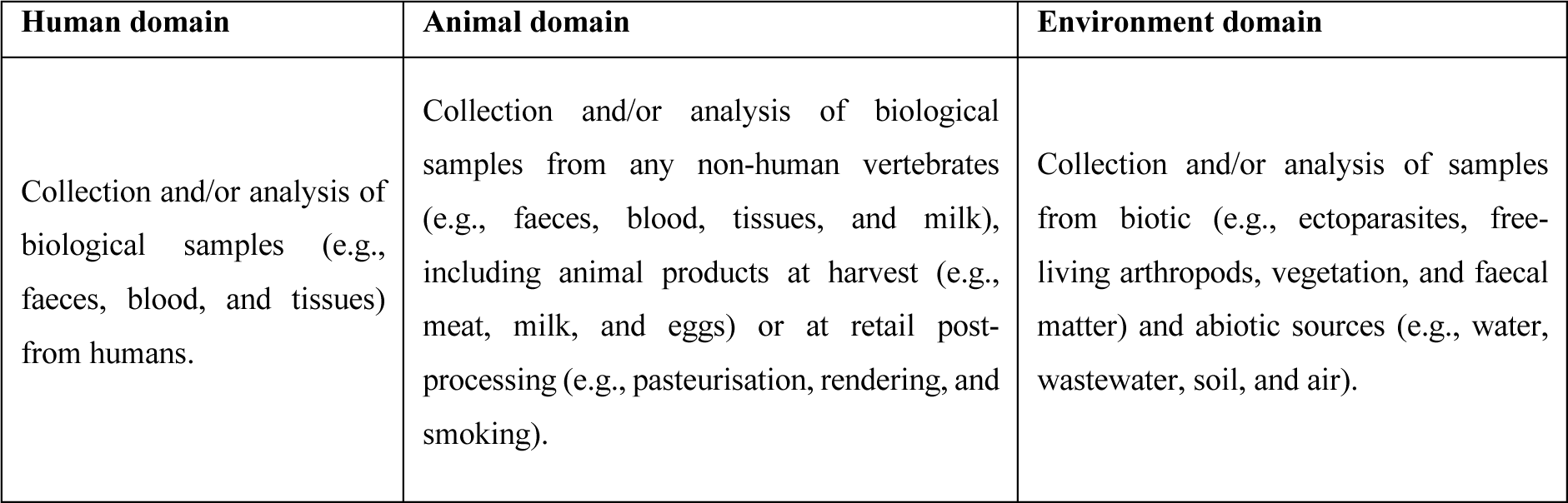
One Health domains. Criteria used to define which One Health domains are covered by each of the 2,992 publications screened as part of our study.

The following studies were excluded from our scoping review: I) scientific work focusing on SARS-CoV-2; II) articles which do not incorporate original genomic data (in other words, we excluded studies that only collated data deposited in publicly accessible databases); III) methodologies exclusively based on Sanger sequencing and amplified/restriction fragment length polymorphism; IV) literature reviews, perspective articles, and commentaries; and V) grey literature and literature whose full text is not available in English.

### Data extraction and analysis

After full-text screening, each retained record was subjected to data extraction to understand its overarching aim, sampling effort, laboratory methodologies, and analytical approach (see Table 3 for full details). We were also interested in reproducibility and accessibility of results, and therefore collated data on whether genetic data and open-source code were submitted to public repositories, and if software used in each study was licenced or open source. Based on the extracted data, geographic localities where sampling was carried were aggregated based on income status (i.e., low/lower-middle income countries and upper-middle/high income countries) as reported by the Organisation for Economic Co-operation and Development in 2022 (https://www.oecd.org/dac/financing-sustainable-development/development-finance-standards/daclist.htm). Furthermore, the biological agents included in our review were categorised based on hazard group definitions by Health and Safety Executive (https://www.hse.gov.uk/pubns/misc208.htm) and current legislation in the UK (i.e., The Specified Animal Pathogens (Scotland) Order 2009 under the Animal Health Act 1981). These categories reflect infectiousness, morbidity, and available vaccines or treatments, which were translated into laboratory containment levels required to work with the listed pathogens.

**Table 3.** Data extraction. Summary of the data extracted from each record included after full-text screening.

| **Publication** | **Description of the recorded data** |
| --- | --- |
| First author's surname | Not applicable |
| Scientific journal | ISO4 abbreviation of the scientific journal and publication's year |
| **Epidemiology** | **Description of the recorded data** |
| Country or geographic region | Geographic origin of the study and where specimens were sequenced |
| Sampling period | Sampling years spanning from the most dated to most recent sample |
| Infectious agent(s) | Pathogen(s) included in the study |
| Study aim(s) | Overarching aim(s) of the study as specified by its Introduction's final paragraph and Conclusions sections |
| One Health domain(s) | Counts of samples from the human, animal, and environment domains which were collected (or analysed) and sequenced by the study |
| Animal domain | Subdivided into livestock (i.e., farmed domestic pigs, ruminants, horses, and fish), pet (i.e., domestic dogs, cats, pet birds, pet rodents, and exotic pets), poultry (i.e., farmed chickens, ducks, geese, guinea fowl, and turkeys), and wildlife (i.e., any non-domesticated vertebrates) |
| Environment domain | Subdivided into abiotic (i.e., water, soil, wastewater, housing/transport/market/slaughterhouse environment, and unspecified environmental samples), Arachnida (i.e., ticks and mites), biotic (i.e., vegetation, animal feed, faecal matter collected from the environment, and unspecified items for human consumptions of non-animal origin), and Insecta (i.e., mosquitoes, flies, fleas, and lice) |
| **Laboratory** | **Description of the recorded data** |
| Isolation by cell culture | Pathogen isolation prior to genome sequencing |
| Detection/Characterisation | Pathogen identification by serological, molecular, or other methods prior to genome sequencing |
| Nucleic acid | Targeted nucleic acid for automated or manual extraction using commercial kits or in-house methods |
| Genome sequencing | Library preparation kits and genome sequencing platform(s) |
| **Data analysis** | **Description of the recorded data** |
| Assembly of reads | Software used for *de novo* assembly or mapping to a reference genome |
| Evolutionary model | Software used to align consensus sequences, identify recombination events, and construct phylogenetic trees |
| Phylogenetic output | Subdivided into genetic distance, phylogenetic relatedness, and temporally resolved models |
| **Data repository** | **Description of the recorded data** |
| Public archive | Format and public repository for the generated genomic data and bioinformatics code |

To understand factors influencing sample size, we constructed a generalised linear model with total sample size of each record (log-transformed with Poisson family links) as the response variable and pathogen type, biocontainment level, sequencing platform, sampling geographic origin, income status stratification, overarching aim of the study, number of surveyed domains, and year of publication as predictors. Data were modelled using quasi-Poisson and negative binomial families and all models were tested for overdispersion. Data were analysed and visualised in R version 4.3.2 (R Core Team, 2023).

## DATA AVAILABILITY

The code used for data analysis and visualisation is available at https://github.com/cfaustus/ngs_scoping_review.git. Enlighten: Research Data, the dataset repository of the University of Glasgow, has been used to make the research data produced by this study available (http://dx.doi.org/10.5525/gla.researchdata.1534).

## Data Availability

https://github.com/cfaustus/ngs_scoping_review.git

## ACKNOWLEDGEMENTS

SC, FB, ZIT, and CJS were sponsored by the Defense Threat Reduction Agency, United States Department of the Defense (HDTRA12110028). The content of the information does not necessarily reflect the position or the policy of the federal government, and no official endorsement should be inferred. CLF was supported by a Natural Environment Research Council Independent Research Fellowship (NE/V01430/1). JR was supported by the UK Research and Innovation Global Challenges Research Fund One Health Poultry Hub (BB/S011269/1).

## AUTHOR CONTRIBUTIONS

**Stefano Catalano:** conceptualization; data curation; investigation; methodology; project administration; writing – original draft; writing – review and editing. **Francesca Battelli:** data curation; writing – original draft; writing – review and editing. **Zoumana I Traore:** data curation; investigation. **Jayna Raghwani:** formal analysis; supervision; visualization. **Christina L Faust:** conceptualization; formal analysis; investigation; supervision; visualization; writing – review and editing. **Claire J Standley:** conceptualization; funding acquisition; investigation; methodology; supervision; writing – review and editing.

