## Supplementary figures and tables for "Pathogen genomics and One Health: a scoping review of current practices in zoonotic disease research"

**Appendix Table S1. Refining the search string using PubMed®.**

Any addition to the initial query string is in bold and underlined whereas deletions are struck through. The final exploratory search hitting a total of 2,083 queries was executed on September 5th 2022.

| Query result | Query string |
|---|---|
| 1,459 | ((ecolog* OR evolution* OR epidemiolog*) AND ("transmission" OR transmit* OR "surveillance") AND (zoono* OR "disease" OR pathogen* OR infect*) AND ("molecular" OR genetic* OR genom* OR metagenom*) AND (phylogen* OR phylodynamic* OR phylogeograph*) AND ("reads" OR librar* OR "mapping" OR align* OR "next generation")) AND ("2005"[Date - Publication] : "3000"[Date - Publication]) |
| 2,497 | ((ecolog* OR evolution* OR epidemiolog*) AND ("transmission" OR transmit* OR "surveillance") AND (zoono* OR "disease" OR pathogen* OR infect*) AND ("molecular" OR genetic* OR genom* OR metagenom*) AND (phylogen* OR phylodynamic* OR phylogeograph*) AND ("reads" OR librar* OR "mapping" OR align* **OR polymorph*** OR "next generation")) AND ("2005"[Date - Publication] : "3000"[Date - Publication]) |
| 2,332 | ((ecolog* OR evolution* OR epidemiolog*) AND ("transmission" OR transmit* OR "surveillance") AND (zoono* OR "disease" OR pathogen* OR infect*) AND ("molecular" OR genetic* OR genom* OR metagenom*) AND (phylogen* OR phylodynamic* OR phylogeograph*) AND ("reads" OR librar* OR "mapping" OR align* **OR polymorph*** OR "next generation") **NOT (Sanger OR microsatellite*)**) AND ("2005"[Date - Publication] : "3000"[Date - Publication]) |
| 2,224 | ((ecolog* OR evolution* OR epidemiolog*) AND ("transmission" OR transmit* OR "surveillance") AND (zoono* OR "disease" ~~**OR pathogen***~~ OR infect*) AND ("molecular" OR genetic* OR genom* OR metagenom*) AND (phylogen* OR phylodynamic* OR phylogeograph*) AND ("reads" OR librar* OR "mapping" OR align* **OR polymorph*** OR "next generation") **NOT (Sanger OR microsatellite*)**) AND ("2005"[Date - Publication] : "3000"[Date - Publication]) |
| 2,103 | ((ecolog* OR evolution* OR epidemiolog*) AND ("transmission" OR transmit* OR "surveillance") AND (zoono* OR "disease" ~~**OR pathogen***~~ OR infect*) AND ("molecular" OR genetic* OR genom* OR metagenom*) AND (phylogen* OR phylodynamic* OR phylogeograph*) AND ("reads" OR librar* ~~**OR "mapping"**~~ OR align* **OR polymorph*** OR "next generation") **NOT (Sanger OR microsatellite*)**) AND ("2005"[Date - Publication] : "3000"[Date - Publication]) |
| 2,083 | ((ecolog* OR evolution* OR epidemiolog*) AND ("transmission" ~~**OR transmit***~~ OR "surveillance") AND (zoono* OR "disease" ~~**OR pathogen***~~ OR infect*) AND ("molecular" OR genetic* OR genom* OR metagenom*) AND (phylogen* OR phylodynamic* OR phylogeograph*) AND ("reads" OR librar* ~~**OR "mapping"**~~ OR align* **OR polymorph*** OR "next generation") **NOT (Sanger OR microsatellite*)**) AND ("2005"[Date - Publication] : "3000"[Date - Publication]) |

**Appendix Table S2. PRISMA-ScR checklist.**

Preferred Reporting Items for Systematic reviews and Meta-Analyses extensions for Scoping Reviews (PRISMA-ScR) checklist adapted to our study.

| Section | Item | PRISMA-ScR checklist item | Reported on page # |
|---|---|---|---|
| **Title** | | | |
| Title | 1 | Identify the report as a scoping review | 1 (Title page) |
| **Abstract** | | | |
| Structured summary | 2 | Provide a structured summary that includes (as applicable): background, objectives, eligibility criteria, sources of evidence, charting methods, results, and conclusions that relate to the review questions and objectives | 2 (Abstract) |
| **Introduction** | | | |
| Rationale | 3 | Describe the rationale for the review in the context of what is already known. Explain why the review questions/objectives lend themselves to a scoping review approach | 3-4 (Introduction) |
| Objectives | 4 | Provide an explicit statement of the questions and objectives being addressed with reference to their key elements (e.g., population or participants, concepts, and context) or other relevant key elements used to conceptualize the review questions and/or objectives | 4 (Introduction's final paragraph) |
| **Methods** | | | |
| Protocol and registration | 5 | Indicate whether a review protocol exists; state if and where it can be accessed (e.g., a Web address); and if available, provide registration information, including the registration number | Not applicable |
| Eligibility criteria | 6 | Specify characteristics of the sources of evidence used as eligibility criteria (e.g., years considered, language, and publication status), and provide a rationale | 12-13 (Methods) |
| Information sources* | 7 | Describe all information sources in the search (e.g., databases with dates of coverage and contact with authors to identify additional sources), as well as the date the most recent search was executed | 12 (Methods) |
| Search | 8 | Present the full electronic search strategy for at least one database, including any limits used, such that it could be repeated | 12 (Methods) and Appendix Table S1 |
| Selection of sources of evidence† | 9 | State the process for selecting sources of evidence (i.e., screening and eligibility) included in the scoping review | 12-13 (Methods) |
| Data charting process‡ | 10 | Describe the methods of charting data from the included sources of evidence (e.g., calibrated forms or forms that have been tested by the team before their use, and whether data charting was done independently or in duplicate) and any processes for obtaining and confirming data from investigators | 13 (Methods) and Table 3 |
| Data items | 11 | List and define all variables for which data were sought and any assumptions and simplifications made | Table 2 |

| | | | |
|---|---|---|---|
| Critical appraisal of individual sources of evidence§ | 12 | If done, provide a rationale for conducting a critical appraisal of included sources of evidence; describe the methods used and how this information was used in any data synthesis (if appropriate) | 12-13 (Methods) |
| Synthesis of results | 13 | Describe the methods of handling and summarizing the data that were charted | 13 (Methods) |
| **Results** | | | |
| Selection of sources of evidence | 14 | Give numbers of sources of evidence screened, assessed for eligibility, and included in the review, with reasons for exclusions at each stage, ideally using a flow diagram | 4 (Results) and Figure EV2 |
| Characteristics of sources of evidence | 15 | For each source of evidence, present characteristics for which data were charted and provide the citations | Table 1 and Figures 1-5 |
| Critical appraisal within sources of evidence | 16 | If done, present data on critical appraisal of included sources of evidence (see item 12) | 4 (Results) |
| Results of individual sources of evidence | 17 | For each included source of evidence, present the relevant data that were charted that relate to the review questions and objectives | Figures 1-5 and Figures EV3-EV4 |
| Synthesis of results | 18 | Summarize and/or present the charting results as they relate to the review questions and objectives | Appendix Tables S4-S6 |
| **Discussion** | | | |
| Summary of evidence | 19 | Summarize the main results (including an overview of concepts, themes, and types of evidence available), link to the review questions and objectives, and consider the relevance to key groups | 7-11 (Discussion) |
| Limitations | 20 | Discuss the limitations of the scoping review process | 9-10 (Discussion) |
| Conclusions | 21 | Provide a general interpretation of the results with respect to the review questions and objectives, as well as potential implications and/or next steps | 11 (Conclusions) |
| **Funding** | | | |
| Funding | 22 | Describe sources of funding for the included sources of evidence, as well as sources of funding for the scoping review. Describe the role of the funders of the scoping review | 14 (Acknowledgements) |

* Where *sources of evidence* (see second footnote) are compiled from, such as bibliographic databases, social media platforms, and Web sites.

† A more inclusive/heterogeneous term used to account for the different types of evidence or data sources (e.g., quantitative and/or qualitative research, expert opinion, and policy documents) that may be eligible in a scoping review as opposed to only studies. This is not to be confused with *information sources* (see first footnote).

‡ The frameworks by Arksey and O'Malley (6) and Levac and colleagues (7) and the Joanna Briggs Institute guidance (4, 5) refer to the process of data extraction in a scoping review as data charting.

§ The process of systematically examining research evidence to assess its validity, results, and relevance before using it to inform a decision. This term is used for items 12 and 19 instead of "risk of bias" (which is more applicable to systematic reviews of interventions) to include and acknowledge the various sources of evidence that may be used in a scoping review (e.g., quantitative and/or qualitative research, expert opinion, and policy document).

**Appendix Table S3. List of pathogens classified as zoonotic.**

List of diseases classified as zoonotic following guidelines of World Health Organization, Public Health England, and European Centre for Disease Prevention and Control. Reference to zoonotic transmission mode is in parentheses. Additional infectious organisms deemed of zoonotic potential during primary and full-text record screenings have been included.

| **Pathogen** | **Disease** | **Zoonotic transmission mode** |
|---|---|---|
| *Acinetobacter baumannii* | Infections in blood, urinary tract, and lungs (pneumonia) | In humans, typically nosocomial transmission in healthcare settings, but possible environmental zoonotic transmission (source) |
| *Alphavirus* genus | Chikungunya and equine encephalitis | Vector-borne diseases spread by mosquitoes (source) |
| *Anaplasma marginale*<br>*A. phagocytophilum* | Anaplasmosis | Vector-borne disease spread by ticks (source) |
| *Avian orthoavulavirus 1* | Newcastle disease | Contact with fluids from infected birds (source) |
| *Babesia* spp. | Babesiosis | Vector-borne disease spread by ticks (source) |
| *Bacillus anthracis* | Anthrax | Environmental contamination of spores, contact with infected animals through handling and consumption (source) |
| *Bacillus cereus* | Gastrointestinal or non-gastrointestinal illness | Food-borne diseases associated with consumption of contaminated soil, meat, milk, vegetables, and fish (source) |
| *Bartonella henselae* | Cat scratch fever | Acquired through scratch of infected domestic or feral cats (source) |
| *Betacoronavirus* genus | Middle East and severe acute respiratory syndrome | Consumption of contaminated animal products and, in some cases, proximity to an infected animal is sufficient; once in humans, it spreads via direct human-to-human contact (source) |
| *Borrelia* spp. | Lyme disease and tick-borne relapsing fever | Vector-borne diseases spread by ticks (source) |
| *Brucella* spp. | Brucellosis | Direct contact with infected animals, consumption of contaminated animal products, inhalation of contaminated aerosols or dust (source) |
| *Burkholderia mallei*<br>*B. pseudomallei* | Glanders and melioidosis | Direct contact with infected animals, inhalation of contaminated aerosols, or dust from infected animals (source) |
| *Campylobacter* spp. | Campylobacteriosis | Food-borne disease likely acquired by consuming raw or undercooked poultry, seafood, meat, produce, and contaminated drinking water (source) |
| *Chlamydia abortus*<br>*C. psittaci* | Ovine chlamydiosis and psittacosis | *C. abortus* in humans is occupationally related, principally occurring by inhalation during sheep births; psittacosis is spread through direct contact |

| | | |
|---|---|---|
| | | with infected birds (source) |
| *Clonorchis sinensis* | Clonorchiasis | Infection in humans is a result of consuming contaminated fish (source) |
| *Clostridium baratii*<br>*C. botulinum* | Botulism | Rare disease in humans, it occurs via consumption of contaminated food (source) |
| *Corynebacterium ulcerans* | Zoonotic diphtheria | Rare disease in humans, it occurs via direct contact with animals or contaminated milk (source) |
| *Cowpox virus* | Cowpox | Transmission via direct contact with infected animals (source) |
| *Coxiella burnetii* | Q fever | Transmission via direct contact with infected animal fluids, inhalation of dust containing bacteria, and ingestion of unpasteurized milk (source) |
| *Crimean-Congo hemorrhagic fever orthonairovirus* | Crimean-Congo haemorrhagic fever | Vector-borne disease spread by ticks or close contact with infected individuals and animals (source) |
| *Cryptosporidium* spp. | Cryptosporidiosis | Ingestion of spores found in soil, food, water, or surfaces contaminated by infected animals (source) |
| *Dabie bandavirus* | Severe fever with thrombocytopenia syndrome | Most likely vector-borne, spread by ticks (source) |
| *Eastern equine encephalitis virus* | Eastern equine encephalitis | Vector-borne disease spread by mosquitoes (source) |
| *Zaire ebolavirus* | Ebola virus disease | Transmission to humans appears to be via direct contact with infectious animals, and direct human-to-human contact (source) |
| *Echinococcus* spp. | Alveolar echinococcosis and hydatid disease | Consumption of contaminated soil, water, or food (source) |
| *Enterobacter cloacae* | Pneumonia, urinary tract infections, and septicaemia | Consumption of contaminated soil, water, or food (source) |
| *Enterococcus faecalis*<br>*E. faecium* | Urinary tract infection, endocarditis, and bacteriaemia | Likely food-borne, mostly through consumptions of contaminated meat (source) |
| *Erysipelothrix rhusiopathiae* | Erysipeloid | *E. rhusiopathiae* in humans is occupationally related, principally occurring after contact with contaminated animals, their products or waste, or soil (source) |
| *Escherichia coli* (shiga toxin-producing) | Haemorrhagic colitis/pneumonia and haemolytic uraemic syndrome | Food-borne disease or directly transmitted through faeces of infected animals (source) |
| *Fasciola gigantica*<br>*F. hepatica* | Fascioliasis | Infection in humans is a result of environmental and water-borne contamination (source) |
| *Flavivirus* genus | Dengue, Kyasanur Forest disease, Japanese/tick-borne encephalitis, West Nile fever, yellow fever, and Zika | Vector-borne diseases spread by mosquitoes or ticks (source) |

| | | |
|---|---|---|
| *Francisella tularensis* | Tularemia | Vector-borne disease spread by arthropods, inhalation of contaminated aerosols, consumption of contaminated water, and contact with infected animals (source) |
| *Giardia* spp. | Giardiasis | Consumption of contaminated food or water, contact with faeces of infected animals (source) |
| *Henipavirus* genus | Angavokely, Hendra, and Nipah virus infection | Transmission to humans appears to be via direct contact with infectious animals (source) |
| *Hepacivirus* genus | Hepatitis C | Transmission to humans remains unclear (source) |
| *Influenza A virus* | Animal/Avian influenza | Transmission to humans appears to be via direct contact with infectious animals (source) |
| Jingmenvirus | Febrile illness, suspected | Likely vector-borne, mainly transmitted to humans through tick bites (source) |
| *Klebsiella pneumoniae* | Pneumonia, wound infection, meningitis, and bloodstream infection | Limited evidence, possible infection through contact with contaminated water or soil and consumption of infected animal products (source) |
| *Kobuvirus* genus | Acute gastroenteritis | Transmission follows the oral route via consumption of contaminated food or water (source) |
| *Lassa mammarenavirus* | Lassa fever | Direct contact with urine or faeces from infected rodents (source) |
| *Leishmania* spp. | Leishmaniasis | Vector-borne disease spread by Phlebotominae flies (source) |
| *Leptospira* spp. | Leptospirosis | Contact with water, soil, or surfaces contaminated with urine from infected animals; consumption of contaminated food or water (source) |
| *Listeria* spp. | Listeriosis | Primarily a food-borne disease with few cases of environmentally acquired bacteria (source) |
| *Louping ill virus* | Louping ill | Rare disease in humans transmitted by ticks or through contact with infected sheep (source) |
| *Lymphocytic choriomeningitis mammarenavirus* | Lymphocytic choriomeningitis | Acquired after contact with excreta of infected rodents (source) |
| *Lyssavirus* genus | Rabies | Transmission to humans appears to be via direct contact with infectious animals or humans (source) |
| *Marburg marburgvirus* | Marburg virus disease | Animal-to-human transmission remains unknown but, once in humans, it spreads via direct human-to-human contact (source) |
| *Monkeypox virus* | Monkeypox | Transmission to humans appears to be via direct contact with infectious animals and humans (source) |

| | | |
|---|---|---|
| *Mycobacterium* spp. | Buruli ulcer, fish tank/swimming pool granuloma, leprosy, paratuberculosis, and tuberculosis | Transmission to humans mainly via direct contact with infectious animals or contaminated animal products (source) |
| *Opistorchis felineus*<br>*O. viverrini* | Opistorchiasis | Food-borne associated with consumption of contaminated fish (source) |
| *Orf virus* | Orf | Transmission to humans seems to be via direct contact (source) |
| *Orientia tsutsugamushi* | Scrub typhus | Vector-borne disease spread by bites of infected chiggers (source) |
| *Orthobornavirus* genus | Borna disease | Contact with excreta of infected animals (source) |
| *Orthohantavirus* genus | Haemorrhagic fever with pulmonary/renal syndrome | Inhalation of faecal aerosols from infected rodents (source) |
| *Orthohepevirus A* (hepatitis E virus) | Hepatitis E | Hepatitis E infections are mostly a result of genotypes 1 and 2 (only found in humans, whereas genotypes 3 and 4 circulate in animals and rarely infect humans); transmission follows the oral route via consumption of contaminated meat (source) |
| *Orthoreovirus* genus | Gastrointestinal, central nervous system, or respiratory symptoms | Direct contact with infected animals and with environmental sources (source) |
| *Paragonimus* spp. | Paragonimiasis | Infection in humans is a result of consuming contaminated crustaceans (source) |
| *Pasteurella* spp. | Pasteurellosis | Direct contact with infected animals or through consumption of contaminated food and water; some evidence for aerosol spread (source) |
| *Pegivirus* genus | Hepatitis G | Transmission to humans remains unclear (source) |
| *Phlebovirus* genus | Rift Valley fever | Vector-borne disease spread by mosquitoes or via direct contact with infected animals (source) |
| *Plasmodium* spp. | Malaria | Vector-borne disease spread by mosquitoes (source) |
| *Pseudomonas aeruginosa* | Blood infections, pneumonia, or localised wound infections | Direct contact with aerosols, saliva, urine, or faeces from infected animals (source) |
| *Rhadinovirus* genus | Kaposi's sarcoma | Transmission to humans remains unclear (source) |
| *Rickettsia* spp. | African tick-bite fever, Mediterranean spotted fever, and Rocky Mountain spotted fever | Vector-borne disease spread by ticks, fleas, and mites (source) |
| Ringworm | Ringworm or dermatophytosis | Fungal disease transmitted via skin contact with infected animals (source) |
| *Rotavirus A/C/G* | Gastrointestinal illness | Direct contact with infected animals and contaminated faeces (source) |
| *Rubivirus* genus | Rubella | Mechanisms of zoonotic transmission remain unclear when infecting humans; the disease spreads via contact with respiratory secretions of infected individuals (source) |

| | | |
|---|---|---|
| *Sapovirus* genus | Acute gastroenteritis | Transmission follows the oral route and food-borne transmission may also occur (source) |
| *Salmonella enterica* | Salmonellosis | Food-borne disease through consumption of contaminated food or contact with infected animals (source) |
| *Schistosoma* spp. | Schistosomiasis | Percutaneous infection after contact with contaminated water (source) |
| *Sindbis virus* | Sindbis fever | Vector-borne disease spread by mosquitoes (source) |
| *Sporothrix* spp. | Sporotrichosis | Fungal disease transmitted via environmental sources or direct contact with infected cats (source) |
| *Staphylococcus aureus* (methicillin-resistant) | Infection of skin, multiple organs, and tissues | Human cases primarily through nosocomial transmission but also through direct contact with infected animals, their products, and fomites (source) |
| *Streptobacillus moniliformis* | Rat bite fever | Rare disease in humans, spread through direct contact with rodents and their excreta (source) |
| *Streptococcus agalactiae*<br>*S. suis*<br>*S. zooepidemicus* | Streptococcal sepsis | Rare disease in humans; most transmission occurs via direct contact with infected animals or meat, with some evidence of airborne transmission (source) |
| *Strongyloides stercoralis* | Strongyloidiasis | Contact with contaminated soil (source) |
| *Taenia* spp. | Cysticercosis/Taeniasis | Food-borne disease associated with consumption of contaminated, under-cooked pork (source) |
| *Toxocara canis*<br>*T. cati* | Toxocariasis | Ingestion of contaminated environmental sources and, in rare cases, meat (source) |
| *Toxoplasma gondii* | Toxoplasmosis | Consumption of contaminated soil, water, or food (source) |
| *Trichinella* spp. | Trichinellosis | Food-borne disease associated with consumption of contaminated, under-cooked meat (source) |
| *Trypanosoma brucei*<br>*T. cruzi* | Sleeping sickness and Chagas disease | Vector-borne diseases spread by arthropod vectors (source) |
| Prion protein | Variant Creutzfeldt-Jakob disease | Presumably food-borne disease via consumption of contaminated beef (source) |
| *Vibrio parahaemolyticus* | Vibriosis | Food-borne disease associated with consumption of contaminated seafood or through direct contact of open wounds with contaminated water (source) |
| *Yersinia enterocolitica*<br>*Y. pestis*<br>*Y. pseudotuberculosis* | Plague, and Far East scarlet-like fever | Transmission via flea bites carried by rodents and, less commonly, via scratches or bites from infected domestic cats, direct handling of infected animal tissues, or inhalation of contaminated particles (source) |

**Appendix Table S4. Zoonotic pathogens and overarching aim(s) of the studies.**

Zoonotic pathogens investigated in the 114 studies included for data extraction and main aim(s) of the studies (i.e., cross-domain transmission (CDT), diagnostics development and surveillance (DDS), genomic diversity and evolution (GDE), genome-wide association studies including antimicrobial resistance (GWA), and outbreak epidemiology investigations (OBE)). The values 1 and 0.5 were assigned when each study had a single main aim or two-fold aims, respectively. The total number of records for each taxon is reported in parentheses.

| **Pathogen** | | **Aims of the study** | | | | |
|---|---|---|---|---|---|---|
| **Classification** | **Taxon** | **CDT** | **DDS** | **GDE** | **GWA** | **OBE** |
| Gram-negative bacteria | *Salmonella enterica* (22) | 3.5 | 2 | 3.5 | 4.5 | 8.5 |
| | *Escherichia coli* (13) | 5.5 | 0 | 0.5 | 6 | 1 |
| | *Burkholderia pseudomallei* (5) | 0 | 0.5 | 3.5 | 0 | 1 |
| | *Campylobacter* spp. (3) | 1 | 0 | 0 | 1.5 | 0.5 |
| | *Francisella tularensis holartica* (3) | 1 | 0 | 1 | 0 | 1 |
| | *Yersinia pestis* (3) | 0 | 0 | 2 | 0 | 1 |
| | *Coxiella burnetii* (2) | 1 | 1 | 0 | 0 | 0 |
| | *Vibrio parahaemolyticus* (2) | 0 | 1 | 0 | 0.5 | 0.5 |
| | *Acinetobacter baumannii* (1) | 0 | 0 | 0 | 0 | 1 |
| | *Chlamydia psittaci* (1) | 1 | 0 | 0 | 0 | 0 |
| | Enterobacteriaceae family (1) | 1 | 0 | 0 | 0 | 0 |
| | *Klebsiella pneumoniae* (1) | 0 | 0 | 0 | 0 | 1 |
| | *Leptospira* spp. (1) | 0 | 0.5 | 0.5 | 0 | 0 |
| | *Orientia tsutsugamushi* (1) | 0 | 0 | 1 | 0 | 0 |
| | *Rickettsia japonica* (1) | 0 | 0 | 1 | 0 | 0 |
| | **Total (60)** | **14** | **5** | **13** | **12.5** | **15.5** |
| Gram-positive bacteria | *Staphylococcus aureus* (11) | 2.5 | 0 | 6.5 | 1 | 1 |
| | *Listeria monocytogenes* (5) | 0 | 1 | 1 | 0 | 3 |
| | *Mycobacterium* spp. (5) | 0 | 0.5 | 4.5 | 0 | 0 |
| | *Bacillus anthracis* (4) | 0 | 1 | 3 | 0 | 0 |
| | *Enterococcus* spp. (3) | 1 | 1 | 0 | 0.5 | 0.5 |
| | *Streptococcus* spp. (3) | 0 | 0 | 2 | 0 | 1 |
| | *Bacillus cereus* (2) | 0.5 | 0 | 0.5 | 0 | 1 |
| | *Clostridium* spp. (2) | 0 | 0 | 0 | 0 | 2 |
| | **Total (35)** | **4** | **3.5** | **17.5** | **1.5** | **8.5** |
| Single-stranded RNA viruses | *Flavivirus* genus (3) | 0 | 0 | 1.5 | 0 | 1.5 |
| | *Influenza A virus* (3) | 0.5 | 0 | 2 | 0 | 0.5 |
| | *Orthohantavirus* genus (3) | 2 | 0 | 1 | 0 | 0 |
| | CCHF *orthonairovirus* (1) | 0 | 0 | 0 | 0 | 1 |
| | *Kobuvirus* genus (1) | 0.5 | 0 | 0.5 | 0 | 0 |
| | *Phlebovirus* genus (1) | 0 | 0 | 1 | 0 | 0 |
| | **Total (12)** | **3** | **0** | **6** | **0** | **3** |
| Others | *Babesia microti* (1) | 0 | 0 | 1 | 0 | 0 |

| | | | | | | |
|---|---|---|---|---|---|---|
| | *Cowpox virus* (1) | 0 | 0 | 0 | 0 | 1 |
| | *Giardia duodenalis* (1) | 1 | 0 | 0 | 0 | 0 |
| | *Plasmodium* spp. (1) | 0 | 0 | 1 | 0 | 0 |
| | *Rotavirus A* (1) | 0 | 0 | 1 | 0 | 0 |
| | *Sporothrix* spp. (1) | 0 | 0 | 1 | 0 | 0 |
| | *Strongyloides stercoralis* (1) | 1 | 0 | 0 | 0 | 0 |
| | **Total (7)** | **2** | **0** | **4** | **0** | **1** |

**Appendix Table S5. Software or pipeline utilised to assemble or map sequencing reads.**

For the 114 studies included for data extraction, we extracted the total number of records (reported in parentheses) detailing software/pipeline for read assembly. Information on *de novo* assembly or mapping to a reference genome was collated when available. The values 1 and 0.5 were assigned when each study applied a single or two analytical approaches, respectively.

| **Software/Pipeline** | ***De novo* assembly** | **Reference mapping** |
|---|---|---|
| SPAdes (31) | 15 | 9.5 |
| CLC Genomics Workbench (15) | 11 | 2 |
| Not specified (12) | 1 | 7 |
| BWA (11) | 0 | 9.5 |
| Velvet (6.5) | 5.5 | 0 |
| Bowtie / Bowtie2 (5.5) | 0 | 5.5 |
| Snippy (5) | 0 | 4 |
| SOAPdenovo (4) | 4 | 0 |
| Newbler (4) | 3 | 0 |
| SMALT (3) | 0 | 3 |
| Geneious (3) | 0 | 2 |
| EDGE Bioinformatics (3) | 0 | 1 |
| Flye (1.5) | 1.5 | 0 |
| MGAP (1) | 0 | 0 |
| SPANDx (1) | 0 | 0 |
| Bionumerics (1) | 0 | 1 |
| SOAP2 (1) | 0 | 1 |
| Platanus (1) | 0 | 1 |
| Qcumber (1) | 0 | 0 |
| Assembler (1) | 0 | 1 |
| A5-miniseq (1) | 0 | 0 |
| Minimap2 (0.5) | 0 | 0.5 |
| Celera Assembler (0.5) | 0 | 0 |
| PEAR (0.5) | 0 | 0 |
| **Total (114)** | **41** | **48** |

**Appendix Table S6. Methods and software utilised for phylogenetic modelling of pathogen genomes.**

For the 114 studies included for data extraction, we extracted the total number of records (reported in parentheses) detailing statistical approaches (i.e., neighbor-joining (NJ), minimum spanning tree (MST), maximum parsimony (MP), maximum likelihood (ML), and Bayesian inference (Bayes)) and software utilised for phylogenetics when available. The values 1, 0.5, and 0.33 were assigned when each study applied a single, two, or three analytical approaches, respectively.

| Software | NJ | MST | MP | ML | Bayes |
|---|---|---|---|---|---|
| RAxML (24.49) | 0 | 0 | 0 | 24.49 | 0 |
| MEGA (15.32) | 6.16 | 0 | 0.33 | 8.83 | 0 |
| BEAST / BEAST2 (10.48) | 0 | 0 | 0 | 0 | 10.48 |
| IQ-TREE (9.16) | 0.5 | 0 | 0 | 8.66 | 0 |
| Not specified (8) | 3 | 2 | 0 | 3 | 0 |
| PhyML (6.99) | 0 | 0 | 0 | 6.99 | 0 |
| FastTree (5.83) | 0 | 0 | 0 | 5.83 | 0 |
| GARLI (4.5) | 0 | 0 | 0 | 4.5 | 0 |
| PAUP* (4.33) | 0 | 0 | 4.33 | 0 | 0 |
| SplitsTree (3.83) | 3.83 | 0 | 0 | 0 | 0 |
| SeqSphere+ (2.83) | 1.5 | 0.33 | 0 | 1 | 0 |
| Bionumerics (2.83) | 1.83 | 0 | 1 | 0 | 0 |
| CSI Phylogeny (2.5) | 0 | 0 | 0 | 2.5 | 0 |
| kSNP3.0 (2) | 1.5 | 0 | 0 | 0.5 | 0 |
| PHYLOViZ (2) | 0 | 2 | 0 | 0 | 0 |
| BactDating (1.5) | 0 | 0 | 0 | 0 | 1.5 |
| Geneious (1.33) | 1 | 0 | 0 | 0.33 | 0 |
| Parsnp (1) | 0 | 0 | 0 | 1 | 0 |
| ApE (1) | 1 | 0 | 0 | 0 | 0 |
| Phangorn (1) | 0 | 0 | 1 | 0 | 0 |
| Gubbins (0.5) | 0 | 0 | 0 | 0.5 | 0 |
| r8s (0.5) | 0 | 0 | 0 | 0.5 | 0 |
| BAPS (0.5) | 0 | 0 | 0 | 0 | 0.5 |
| TreeTime (0.5) | 0 | 0 | 0 | 0 | 0.5 |
| Enterobase (0.33) | 0 | 0 | 0 | 0.33 | 0 |
| TCS (0.33) | 0 | 0 | 0.33 | 0 | 0 |
| GrapeTree (0.3) | 0 | 0.33 | 0 | 0 | 0 |
| **Total (114)** | **20.3** | **4.7** | **7** | **69** | **13** |

**Appendix Figure S1. Decision tree.**

Decision tree representing the inclusion (IC) and exclusion (EC) criteria applied during both title/abstract and full-text screenings of records.

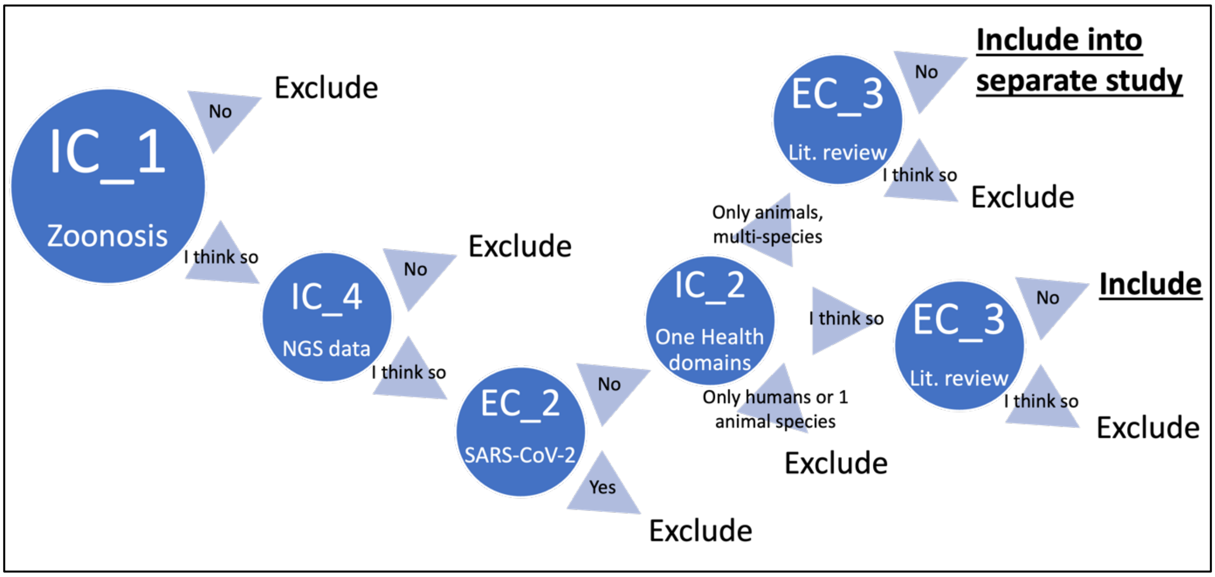

**Appendix Figure S2. PRISMA flow diagram.**

Preferred Reporting Items for Systematic Reviews and Meta-Analyses (PRISMA) flow diagram detailing the number of records retained and excluded at each stage of the screening process.

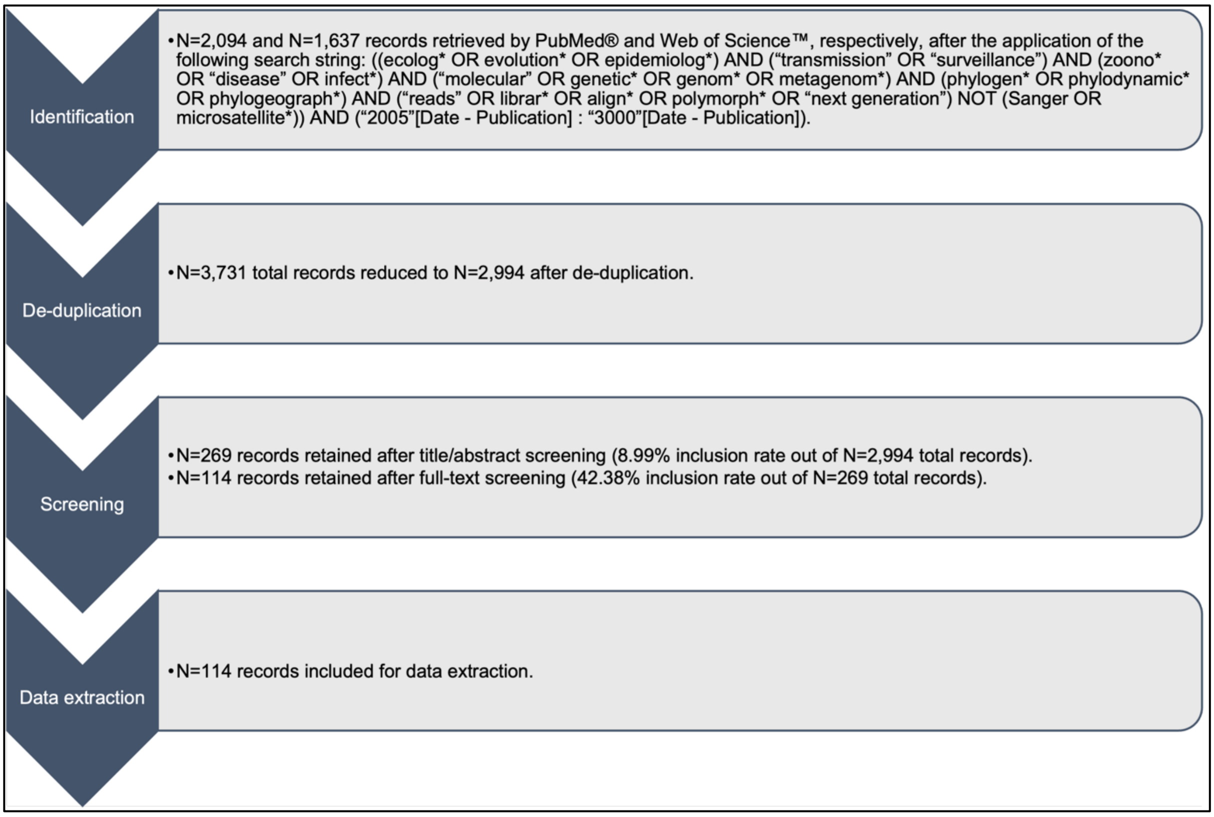

**Appendix Figure S3. Counts of specimens collected (or analysed) and sequenced by each study shaded by its overarching aim*.**

A Histograms representing the sample size reported by each publication (x-axis) and the number of studies (y-axis) for each One Health domain (i.e., human, animal (subdivided into livestock, pet, poultry, and wildlife), and environment (subdivided into abiotic, Arachnida, biotic, and Insecta)) from which the analysed specimens originated.

B Histogram representing the sample size reported by each publication (x-axis) and the number of studies (y-axis).

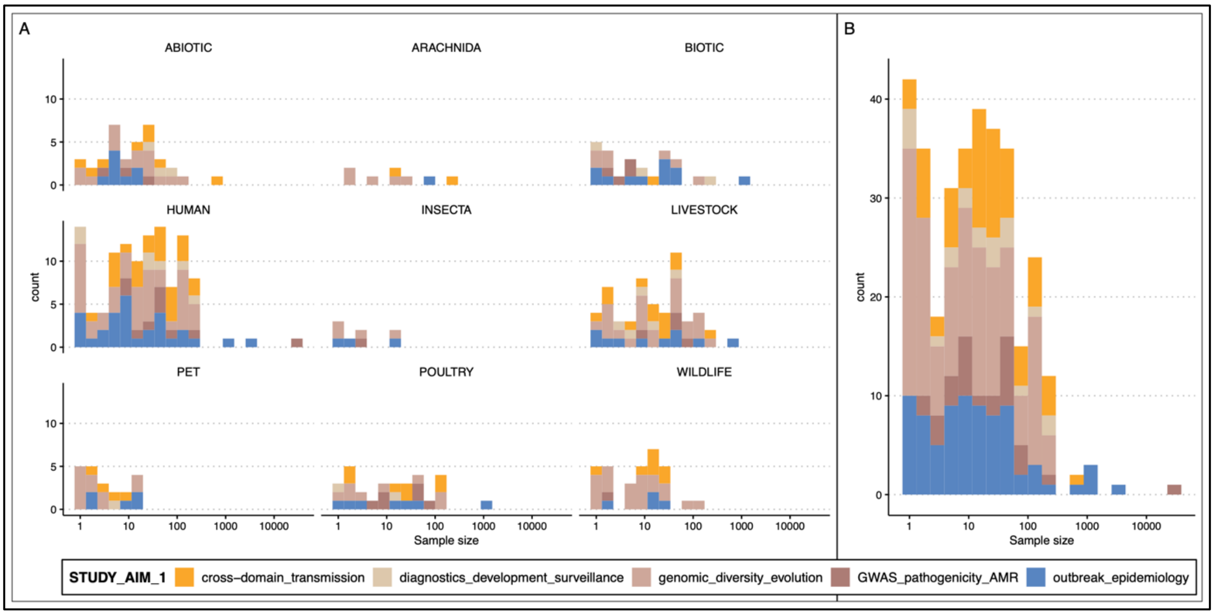


* Cross-domain transmission, diagnostics development and surveillance, genomic diversity and evolution, genome-wide association studies (GWAS) including antimicrobial resistance (AMR), and outbreak epidemiology investigations.

**Appendix Figure S4. Patterns of reproducible science by geographies.**

The alluvial plot shows the proportion of studies that used open science in each of three categories: software, data, and code. A true status means that the publication used software that is open-source or freely available, deposited genomic data and code in publicly accessible repositories. The alluvial flows show the relative abundance of studies that transition between categories and are coloured by geographic origin. Geographic localities where sampling was carried were aggregated based on income status (i.e., low/lower-middle income countries (lmic), upper-middle/high income countries (hic), or both).

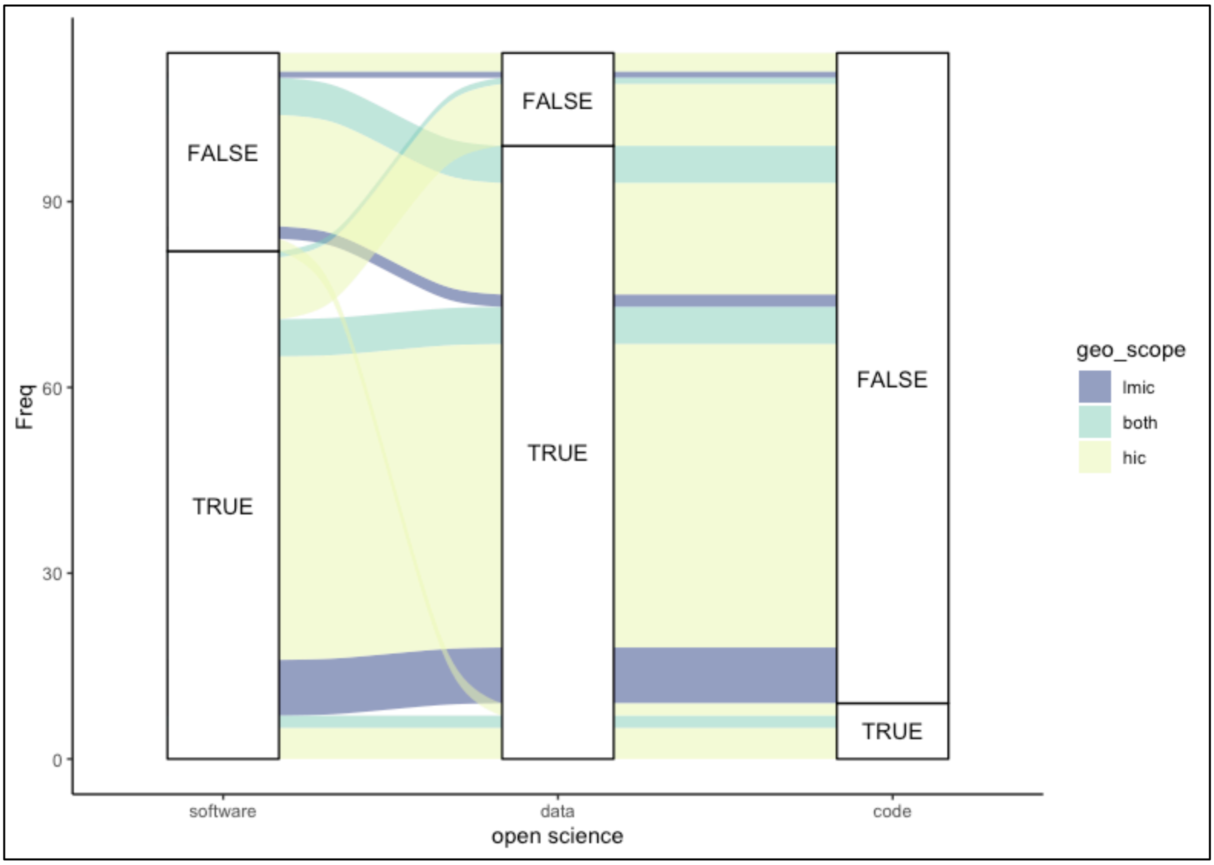
